# Supporting continuous glucose monitoring for people with serious mental illness and type 2 diabetes: Protocol for a co-design study

**DOI:** 10.1101/2024.05.16.24307473

**Authors:** Jennifer VE Brown, Ramzi Ajjan, Najma Siddiqi, Ian Kellar, Peter A Coventry

**Author notes:** **Corresponding author:** Jennifer VE Brown, Mental Health & Addiction Research Group, Department of Health Sciences, University of York, York, UK.

## Abstract

**Introduction:** Compared with the general population, people with serious mental illness (SMI) are 2-3 times more likely to develop type 2 diabetes, have poorer outcomes, and die 15 to 20 years younger, often as a result of long-term physical health conditions. Standard diabetes care does not meet the needs of people with SMI and they are frequently excluded from research, missing out on innovation. As diabetes care increasingly uses technology like continuous glucose monitoring (CGM) it is important to consider the views of people with SMI when new interventions are developed. This is a study protocol to identify candidate components of a structured CGM intervention for people with SMI, including the co-design of a logic model and programme theory.

**Methods:** Drawing on experience-based co-design (EBCD) methods, we propose to collaborate with service-users, carers, and healthcare professionals to undertake early-phase development work for a novel intervention that maximises the potential of CGM to facilitate behaviour change. Fifteen participants will be recruited through existing cohorts and networks in England. The co-design will be informed by existing evidence and based on links between mechanisms of action and behaviour change techniques. Through a series of events (discovery sessions, co-design workshop, celebration event), we will identify candidate components for a prototype intervention ready for further development and testing. A logic model and programme theory will be developed and refined iteratively.

**Discussion:** The main output of this study will be a logic model and programme theory for a novel prototype intervention, ready for further testing following best practice intervention development, such as the Medical Research Council guidance for the development and evaluation of complex interventions. An intervention that makes CGM accessible for people with SMI has the potential to make a considerable contribution to reducing the profound health inequalities experienced by this population.

## Introduction

### Serious mental illness and type 2 diabetes

People with serious mental illness (SMI), conditions like schizophrenia, bipolar disorder, or psychosis, are two to three times more likely to have type 2 diabetes than the general population.(1, 2, 3) The risk of diabetes complications and poorer outcomes is also increased.(4) Diabetes, alongside other long-term physical health conditions, contributes to a reduction in life expectancy by about 15 to 20 years.(5) While this profound example of health inequality is well recognised internationally and is often called the “mortality gap”, attempts to “close the gap” have so far had limited success.(6) People with SMI continue to be less healthy and die earlier than the general population, with most early deaths caused by poor physical health, despite a growing number of research and policy initiatives aimed at reducing health inequalities among this population.

Additionally, people with SMI often miss out on healthcare innovation because they tend to be excluded from research studies.(7) As a group that engages differently with the healthcare system than the general population and frequently requires additional support to successfully access care,(8) people with SMI often do not benefit from existing interventions and services when they are offered without necessary adaptations.(9)

While self-management interventions for mental health have been found to be moderately effective,(10) diabetes self-management skills are often poorly developed among people with SMI; diabetes self-management is perceived as challenging,(11) and existing education programmes like DESMOND(12) do not address the additional needs arising from the dual challenge of managing diabetes alongside SMI.(13, 14)

### Advances in diabetes care through continuous glucose monitoring

Technology plays an increasingly prominent role in diabetes care worldwide. Advances including continuous glucose monitoring (CGM) have introduced lasting changes not only to the delivery of care but also to the individual experience for people with diabetes.(15, 16, 17, 18, 19) CGM devices are two-part systems consisting of a body-worn sensor and an external reader (see Bruttomesso et al.(20) and Lin et al.(21) for an overview of available systems). CGM was introduced to simplify regular blood glucose monitoring. Recent systematic reviews confirm moderate levels of effectiveness of CGM in lowering glycated haemoglobin (HbA1c) in individuals with type 2 diabetes(22) and those with type 1 diabetes.(23)

However, evidence of effectiveness is not observed universally.(24) A crucial discovery has been that glucose monitoring alone does not automatically lead to an improvement in diabetes management and a reduction in HbA1c.(24, 25) It is important to consider the context within which CGM is used. Glucose monitoring should be viewed as an intervention in itself, rather than merely a data collection tool.(26) Deployed on its own, i.e. without support and education for healthcare professionals (HCPs), the person with diabetes, and their carer(s), glucose monitoring is likely to be a waste of resource and studies continue to fail to show evidence of clinical effectiveness defined as a reduction in blood glucose.(27) There is growing evidence that structured glucose monitoring that includes “wrap-around” support, education, and feedback has the potential to improve diabetes management, lower HbA1c, and improve wider health and wellbeing outcomes, including quality of life,(28, 29) pointing towards a synergistic effect of combining glucose monitoring with education and support.

While CGM is being rolled out to a growing population of children and adults with diabetes, people with SMI are not given special consideration,(30, 31) despite the potential of these devices to unlock engagement with the condition and the need to self-manage.(32) In the UK, people with SMI are offered an annual measurement of HbA1c as part of a wider physical health check and medication review in primary care.(33) However, beyond this annual check, people with SMI often struggle to access specialist diabetes care,(34) reinforcing inequities in the provision of care compared with the general population.

### Including people with SMI in the future of CGM research

Further research is warranted into how CGM might improve diabetes self-management and, in turn, glucose management and overall health and wellbeing, for people with SMI. This area is under-researched and the potential of CGM to facilitate behaviour change in this disadvantaged population remains untapped. Work is ongoing to test the effectiveness of a new co-designed supported diabetes self-management intervention for people with SMI.(35, 36, 37) However, while the DIAMONDS feasibility study included a test of the acceptability of CGM in a sample of 21 people with SMI, the sensors were not part of the intervention.

Our previous research has produced indicative findings that wearing a CGM sensor might be acceptable to people with SMI and that they may have an interest in seeing their data, which could be an initial step toward engaging with self-management. Qualitative findings suggest that increased awareness of their diabetes and the CGM sensor acting as a prompt might be key mechanisms that could be exploited as pathways to behaviour and lifestyle changes.

From previous work we also know that support with fitting the device as well as with the interpretation of the data will be crucial to maximise the chances of success. Similarly, our findings suggest that HCPs involved in the care of people with SMI and diabetes will require tailored education depending on their professional background and expertise.

With the exception of SMI-specific studies, people with SMI continue to be largely excluded from health research, either explicitly through restricted eligibility criteria or implicitly through the use of methods that are inaccessible to this population.(7, 38, 39) For a vulnerable group with a disproportionately high illness burden who engage differently with healthcare than the general population and who do not tend to benefit from mainstream education and self-management interventions, this is clearly problematic. This study builds on our own findings as well as the limited existing evidence base about self-management of physical long-term conditions among people with SMI to take a more tailored approach that considers the needs of people with SMI. In this study, we propose to co-design a logic model and programme theory with service-users, carers, and HCPs for a structured CGM intervention for people with SMI.

### Theoretical foundations and guiding frameworks

In line with best practice for the efficient design of evidence-based behaviour-change interventions,(40, 41) the intervention development process will be informed by existing evidence about mechanisms of action (MoAs)(42) and behaviour change techniques (BCTs)(43) that have been shown to be effective in self-management interventions. The Theory and Techniques Tool offers a systematic approach to linking MoAs and BCTs,(44) an approach that has previously been employed in the context of SMI and diabetes self-management as shown in the development of the DIAMONDS intervention.(37)

This co-design study is situated within a context of existing interventions, including CGM itself and the DIAMONDS self-management intervention. However, the ambition is to develop a logic model and programme theory for a new, bespoke intervention that addresses specifically the lack of CGM support and education that has been co-designed and is accessible to people with SMI. As such, this study sets out to co-design a novel intervention rather than an adapt an existing one.(45) Broadly, we will follow the Medical Research Council guidance for the development and evaluation of complex interventions(40) whilst also drawing on principles of multiphase optimisation strategy in determining candidate intervention components ready for further testing.(46)

This protocol is reported in line with GUIDED (GUIDance for the rEporting of intervention Development),(47) with a completed checklist included in Appendix 1.

### Target population, study aim, and objectives

The target population for the new intervention are adults (aged 18 years or over) living in the community with confirmed diagnoses of both type 2 diabetes and SMI (schizophrenia, schizoaffective disorder, psychosis, bipolar disorder, severe depression).

Building on existing evidence in the general population, and feasibility testing and qualitative research that suggests acceptability of CGM in people with SMI, we propose to co-design with service users, carers, and healthcare professionals (HCPs) a logic model and programme theory that include candidate components for a structured CGM intervention. Specifically, we will address the following objectives:

1. To identify candidate intervention components from existing evidence, including the identification of BCTs and MoAs
2. To establish consensus on candidate components of structured CGM from service users and carers
3. To establish consensus on candidate components of structured CGM from HCPs
4. To synthesise outputs from objectives 1 to 3 to develop a preliminary logic model and programme theory

It is not within the scope of this study to include technical innovations in the refinement or further development of CGM devices or systems.

## Methods

We will undertake an accelerated co-design study of candidate intervention components, drawing on established experience-based co-design (EBCD) methods(48) as well as other knowledge translation strategies that have been shown to support co-design.(49) The project will involve the following steps:

1. Synthesis of existing evidence
2. Discovery session with service-users and carers
3. Discovery session with HCPs
4. Reflection and synthesis
5. Joint co-design event with breakout groups, including service-users, carers, and HCPs
6. Synthesis and prototyping
7. Joint showcase and celebration, including service-users, carers, and HCPs

### The project team

The project will be led by JB, a behaviour change researcher with a background in health psychology and experience in applied health research. JB will be supported by PC, RA, IK, and NS who are senior researchers with expertise in health services research, diabetes, health psychology, and psychiatry, respectively. In addition, a member of an established service-user and carer group will be a part of the project team, contributing lived experience of SMI and diabetes. The project will be supported by a trained independent facilitator.(50)

### Participants

We will recruit three groups of participants to take part in this study: service-users, carers, and HCPs. Our aim is to have equal representation of each group, and we will strive to recruit five service-users, five carers, and five HCPs (total N=15). Service-users and carers will be recruited from existing cohorts of participants from previous studies who have given consent to be recontacted about future research opportunities. In addition, participating service-users will be asked to identify informal carers who may be interested in taking part. We are using the King’s Fund definition(51) of informal carers which has been adopted by the NHS and Department of Health and Social Care and includes anyone looking after “*a relative or friend who needs support because of … illness, including mental illness*”, offering “*active support, supervision, or social interaction*”. HCPs will be recruited from existing networks and contacts, using a snowballing approach to building a diverse sample that includes practitioners with expertise in mental health care, such as psychiatrists or mental health nurses, as well as diabetes care, like diabetologists, endocrinologists, or specialist nurses. In addition, we will recruit primary care professionals, i.e. general practitioners (GPs) or practice nurses. We will not restrict participation by role, job title, or seniority. Appendix 2 describes the recruitment and consent process in detail, including examples of the participant-facing documents.

### Eligibility criteria

Table 1 summarises the eligibility criteria for the three groups of participants described above.

**Table 1:** Eligibility criteria for participation.

|  | Inclusion | Exclusion |
| --- | --- | --- |
| **All participants** |  |  |
| *Age* | 18 years or older | Under 18 years |
| *Capacity* | Capacity to consent | No capacity to consent |
| *Cognitive impairment* | No or mild cognitive impairments not affecting ability to provide informed consent. | Any cognitive impairment that affects ability to provide informed consent. |
| *Language<sup>1</sup>* | Ability to communicate in spoken English. | No or severely limited ability to communicate in spoken English, e.g. requiring the use of an interpreter. |
| *Time commitment<sup>2</sup>* | Commitment to attending and contributing to at least two out of three sessions (discovery session, co-design workshop, celebration event). | Not able to commit to at least two out of three sessions. |
| **Additional criteria for service-users** |  |  |
| *SMI diagnosis* | At least 1 of the following<br>Schizophrenia<br>Schizoaffective disorder<br>Bipolar disorder<br>Psychosis | Any other mental health diagnosis, regardless of severity or duration.<br>No mental health diagnosis. |
|  | Severe depression<br>Patients with or without other co-occurring mental health conditions are eligible. |  |
| *Diabetes diagnosis* | Type 2 diabetes (insulin-treated or non-insulin-treated).<br>Patients with or without other co-occurring physical health conditions are eligible. | Any other type of diabetes, including<br>Gestational diabetes<br>Type 1 diabetes<br>Diabetes secondary to other health conditions, e.g. genetic defects |
| *Living situation* | Community dwelling<br>Living in supported housing or residential settings with some degree of autonomy over day-to-day activities | Current inpatients |
| **Additional criteria for carers** |  |  |
| *Caring responsibilities* | Current or recent unpaid carer of a person meeting the criteria outlined above for service-user participants, e.g. a family member, spouse, or friend.<br>Carers are eligible to take part regardless of whether the person they care for (service-user) is taking part in the study. | Paid, i.e. professional, carers.<br>Family members, spouses, or friends of service-users who do not provide care. |
| **Additional criteria for HCPs** |  |  |
| *HCP role and experience* | Any HCP, regardless of job role or seniority, with current or recent experience in at least one of the following:<br>Primary care, e.g. annual physical health checks for people with SMI<br>Mental health care delivered in community-based secondary care settings for people with SMI, e.g. clozapine clinics, community mental health teams, early intervention psychosis clinics<br>Diabetes care delivered in secondary care settings, e.g. diabetes clinics | Any other HCP, including<br>Mental health professionals without direct experience working with people with SMI<br>Mental health professional working exclusively with inpatients<br>HCPs from any specialty not relevant to mental health or diabetes. |

<sup>1</sup>*Language:* We will not formally assess prospective participants' level of English, but a sufficient grasp of the language is crucial for participation in this study. Where possible, we will make adjustments, for example by reading aloud study information materials and limiting the amount of reading and writing participants are asked to do.
<sup>2</sup>*Time commitment:* Participants will have the right to stop their participation at any point and without having to provide a reason. However, at the point of recruitment, prospective participants will be informed that participation involves attendance at three events, and they will be encouraged to commit to attending at least
two out of the three. This is to ensure continuity of participants across the study and to minimise the need for recruitment of additional participants partway through the project. We will aim to over-recruit participants from all three groups to mitigate against the risk of low attendance.

### Project plan

#### Step 1: Synthesis of existing evidence

Findings from a recent mixed-methods systematic review of the acceptability and feasibility of CGM in people with diabetes(52) that have been analysed within the Theoretical Framework of Acceptability (TFA)(53) will be interrogated further using the Theory and Techniques Tool linking BCTs and associated MoAs (i.e., short-term outcomes). In particular, we will extract detail from included studies that describe a structured CGM intervention, i.e. the use of “wrap-around” education and support. In addition, we will draw on findings from a mixed-methods feasibility study of using CGM in a population of people with type 2 diabetes and SMI which have also been analysed and synthesised using the TFA. This will include a description of the CGM systems available in the UK as well as an explanation of different ways of accessing glucose data.

We will complement this evidence with key findings from other programmes of research looking to improve the physical health of people with SMI, particularly those that include the use of technology in their interventions, e.g. DIAMONDS(37) and Stepwise.(54)

To mobilise the synthesised evidence base for the participants in our study, we will develop an animated catalyst film, a number of fictional personas, and descriptions of MoAs and BCTs in accessible language, contextualised for use in the discovery sessions and co-design event.

#### Steps 2 & 3: Discovery sessions

We will conduct separate discovery sessions with different groups of participants. Independently, people with SMI and type 2 diabetes and their informal carers (group 1) and HCPs (group 2) will be asked to use existing evidence to stimulate rapid idea generation for candidate intervention components and content:

1. **Catalyst film**: This short animation will present fictional characters in situations that are likely to be familiar to participants, e.g. challenges in navigating the healthcare system or difficulties engaging with physical health self-management
2. **Personas**: The characters from the catalyst film will additionally be presented as personas that are designed to resonate with participants and their own experiences.
3. **Journey mapping/pain point analysis**: We will work with participants to understand the key barriers (“pain points”) in the patient journey using the personas, i.e. their pathway through the healthcare system within which they are seeking support.
4. **Prototyping:** We will provide accessible minimal viable prototype paper-based BCTs for participants to cluster to pain points and comment on.

Throughout the session, materials will be presented to participants to facilitate discussion and the generation of ideas. Participants will be encouraged to go beyond, add to, and contradict the presented materials. In the synthesis sessions, new content provided by participants will be prioritised. All ideas generated in the discovery sessions will be captured through note taking during discussions as well as photographs of any written outputs, such as flipcharts or sticky notes.

#### Step 4: Reflection and synthesis

Outputs from the discovery sessions relating to the acceptability of CGM will be coded against the TFA for further discussion in step 5 (joint co-design session). Those outputs that relate more directly to BCTs and/or MoAs will be integrated with the initial mapping framework that was produced in step 1.

The main objective of the synthesis and reflection phase will be to produce expanded examples of possible intervention components, highlighting the links between BCTs and MoAs. This will be informed by the evidence synthesised in step 1 as well as outputs from steps 2 and 3.

To support knowledge mobilisation, at this stage we will develop an accessible logic model in lay language alongside a logic model that uses the formal BCT/MoA language which will illustrate putative mechanisms between potential intervention components while also capturing the role of the context within which the intervention is likely to be situated.

#### Step 5: Joint co-design event with breakout groups

For this session, all participants (service-users, carers, and HCPs) will work together with the research team. The focus will be on reaching consensus about key BCTs through joint prioritisation.

Mirroring the approach used in steps 2 and 3, some materials will be presented to participants to enable informed and concentrated conversations. As before, participants will be encouraged to expand on and move beyond these initial prompts.

Initially, participants will be presented with the outputs from the reflection and synthesis stage, i.e. examples of BCT/MoA links and how they might fit within a draft logic model. There will be a chance for participants to add further BCTs (intervention components) that they feel are missing. The final list will then be subject to a prioritisation exercise whereby participants will work individually or in small groups (depending on preference) to rank the BCTs in order of importance.

The most important BCTs will be developed further, e.g. through sketching, prototyping, or in the form of written comments.

#### Step 6: Synthesis and prototyping

As previously, the research team will meet after the co-design event to summarise and synthesise outputs. At this stage, the focus will be on the integration of the expanded BCT/MoAs links from step 5 into the draft logic model and programme theory. The aim of this synthesis stage is to produce a plain language, easily accessible description of what a structured CGM intervention may look like.

#### Step 7: Joint showcase and celebration

The focus of this final event will be to share the intervention prototype and to acknowledge and celebrate the contributions of all participants in the development process. After the initial presentation of the prototype, there will be a chance for further discussion and feedback, this will be free flowing rather than the structured workshop format of the previous sessions. Any pertinent points will be recorded and considered for inclusion for the next stages after completion of this study.

### Decision making priorities and guiding principles

Throughout the project, we will aim to prioritise service-user and carer views and perspectives. However, we acknowledge the need for pragmatism and compromise while operating within time and funding constraints. To ensure that the amount of session content is manageable, reflection and synthesis meetings will be used to prioritise items for discussion. Where there are differences of opinion between the three groups of participants, we will aim to explore those in discussion at the subsequent sessions to reach a deeper understanding and, wherever possible, a mutually acceptable compromise.

Disagreements between participant preferences and published evidence will be addressed in a collaborative fashion with the aim of finding a way to include the published evidence in the intervention in such a way that is acceptable to service users, carers, and HCPs. Conflicting ideas can also be captured in the design of the candidate intervention components and explored in future evaluation.

## Responsible research and innovation (RRI)

Appendix 3 summarises our consideration of equity, diversity, and inclusion in this project as well as reflections on expected participant burden and our plans for participant recognition.

### Study management

Information about study management, governance, and data protection, as well as participant and researcher safety and wellbeing can be found in Appendix 4.

## Knowledge translation and pathways to impact

The main output from this study will be a programme theory and logic model for a structured CGM intervention for people with SMI ready for further testing and refinement. Building on the programme theory, the next step will be to develop a co-designed intervention that addresses the needs of service-users and carers, including training materials for facilitators. This intervention will then need to be further refined and optimised through testing of acceptability and feasibility prior to definitive evaluation.

We will work in an iterative, rather than linear, fashion to avoid premature progression to efficacy and effectiveness testing if further intervention development or refinement work is needed. If the composition of the new intervention allows, we will consider a multi-phase optimisation strategy (MOST) approach which would enable us to understand the contribution of each intervention component as well as any interactions between them.(55) We will continue to collaborate with service users, carers, as well as HCPs to refine the logic model for the new intervention which will inform the design of further studies.

## Discussion

The proposed study will draw together evidence about the acceptability and feasibility of CGM in the general population with diabetes, about diabetes self-management among people with SMI, and emerging findings about the ways CGM might influence behaviour. Guided by established links between MoAs and BCTs, as per the Theory and Techniques tool, we will work with service-users, carers, and HCPs to build on these foundations and co-design a programme theory and logic model for a structured CGM intervention. The study will take a collaborative approach throughout, and service user and carer perspectives will be foregrounded.

The proposed study is situated within a context where we know little about CGM use, uptake, or effectiveness among people with SMI. A recent trial-based cohort study of 279 adults with type 2 diabetes in Australia using CGM, revealed a 1.4% prevalence of schizophrenia/bipolar disorder (4/279 participants). Understandably, due to the very small numbers, no correlation was reported between glycaemic measures, including HbA1c and time-in-range, and SMI.(56) In CGM research as in diabetes care the specific focus on SMI is often lacking and considerations of mental health rarely extend beyond common mental disorders or diabetes distress. However, the importance of behavioural and contextual factors is recognised increasingly as crucial in shaping the future of diabetes care, especially for type 2 diabetes.(57) For example, Hermanns et al. (2022)(58) argue that the integration of behavioural, psychological, and glycaemic data could be key in the drive towards precision medicine in diabetes care by supporting personalised and real-time treatment adjustments and changes to care. They suggest that this might also be true for people with bipolar disorder or depression whereby monitoring could predict relapses which could be avoided with early intervention, consequently leading to an avoidance of deteriorating glucose values. Whether or not this is feasible in practice is unclear at this stage. Firstly, changes in physical activity may be difficult to detect among people with SMI who tend to have extremely sedentary lifestyles.(59) Secondly, people with SMI may not consent to the intensive ongoing monitoring and the use of Artificial Intelligence for this kind of precision-focused approach.(60)

Innovative work by Lee et al. (2023)(61) highlights the challenges that arise from the growing complexity when different data sources, types, and measurements are combined to deliver more personalised diabetes care. In their three-arm RCT they compared treatment as usual (group A), with use of a digital integrated healthcare platform (group B), and with use of the data platform, feedback from HCPs, and intermittent CGM (group C). Both groups B and C showed lower HbA1c and greater weight loss at follow-up compared with group A. Group C showed greater improvements than group B, but these did not reach statistical significance. The intervention received by group C was so complex that it is impossible to unpick from the outcome data alone which component(s) made the difference beyond the data platform and, indeed, if individual components acted antagonistically and cancelled each other out.

The current state of the evidence supports the notion that targeted work in collaboration with people with SMI is very much needed.(57) In a commentary on Lee et al., Kahkoska and colleagues (2023)(62) propose four priorities to maximise the potential of technology-enhanced behaviour-change interventions that are applicable across the population of people with type 2 diabetes, including those who also have SMI. Kahkoska et al. suggest that equity, personalisation, integration with health services, and rigorous evaluation will be crucial going forward. The work proposed here very much aligns with these priorities.

The new intervention will have the potential to address profound health inequalities by improving diabetes care for people with SMI who face higher risks, poorer outcomes, and reduced life expectancy. Current care is clearly not meeting the needs of this vulnerable population; making CGM accessible in a scalable fashion has tremendous potential to improve the lives of some of the most disadvantaged in society. Evidence from a cohort of 813 children with type 1 diabetes suggest that CGM partially mediated the considerable inequalities that are observed in correlations between socioeconomic status and glycaemic management, where those from more deprived areas tend to show poorer management.(63) Exploring the potential of CGM to close, or at least narrow, the mortality gap is imperative and there is no reason to assume that the same mediating effect might not be observed among adults with type 2 diabetes, where similar patterns of inequalities are evident.(64, 65)

## Conclusions

There is a clear need for the development of an intervention that supports people with SMI to benefit from CGM. Co-design has been shown to produce highly acceptable interventions and upon development of a programme theory and logic model we expect to be in a strong position to develop a novel intervention that has the potential to address profound inequalities.

## Data Availability

No datasets were generated or analysed during the current study. All relevant data from this study will be made available upon study completion.

